# High levels of SARS-CoV-2 specific T-cells with restricted functionality in patients with severe course of COVID-19

**DOI:** 10.1101/2020.07.08.20148718

**Authors:** David Schub, Verena Klemis, Sophie Schneitler, Janine Mihm, Philipp M. Lepper, Heinrike Wilkens, Robert Bals, Hermann Eichler, Barbara C. Gärtner, Sören L. Becker, Urban Sester, Martina Sester, Tina Schmidt

## Abstract

Patients infected with SARS-CoV-2 differ in the severity of disease. In this study, SARS-CoV-2 specific T-cells and antibodies were characterized in patients with different COVID-19 related disease severity. Despite severe lymphopenia affecting all major lymphocyte subpopulations, patients with severe disease mounted significantly higher levels of SARS-CoV-2 specific T-cells as compared to convalescent individuals. SARS-CoV-2 specific CD4 T-cells dominated over CD8 T-cells and closely correlated with the number of plasmablasts and SARS-CoV-2 specific IgA- and IgG-levels. Unlike in convalescents, SARS-CoV-2 specific T-cells in patients with severe disease showed marked alterations in phenotypical and functional properties, which also extended to CD4 and CD8 T-cells in general. Given the strong induction of specific immunity to control viral replication in patients with severe disease, the functionally altered phenotype may result from the need for contraction of specific and general immunity to counteract excessive immunopathology in the lung.

## Introduction

Coronavirus disease 19 (COVID-19), the disease caused by the severe acute respiratory syndrome coronavirus 2 (SARS-CoV-2) can be asymptomatic or mild, but also includes severe disease manifestations such as acute respiratory distress syndrome (ARDS), which can lead to multi-organ failure and death despite intensive medical treatment. The mortality rate is particularly high in older individuals and in patients with pre-existing lung, heart, or immunodeficiency diseases^1, 2^.

First studies have shown that SARS-CoV-2 infection causes global changes in cellular immunity, mainly characterized by lymphopenia, skewed distribution of T-cell subpopulations and high plasma concentrations of pro-inflammatory cytokines^2, 3^. In addition, T-cell functionality appears to be altered as shown by impaired expression of interferon gamma (IFNγ)^4-6^. So far, mainly non-specific general changes in the number and functionality of blood cells have been described, whereas specific T-cell immunity directed against SARS-CoV-2 is as yet poorly characterized^7, 8^, especially in patients with different disease severity.

It seems reasonable to suggest that the individual course of a SARS-CoV-2 infection depends on the induction and functionality of the adaptive immunity including both antibodies and T-cells. Seroconversion in patients with COVID-19 does not seem to be delayed, as SARS-CoV-2 specific IgM and IgA antibodies are induced early after the onset of symptoms after a median of 5 days, while the median time for IgG seroconversion is 14 days^9-11^. Thus far, it remains to be elucidated whether patients with different disease manifestations differ in the levels and functionality of SARS-CoV-2 specific T-cells or antibodies. We have previously shown that symptomatic infections with persistent pathogens are associated with alterations in pathogen-specific T-cell levels and impaired functionality as compared to individuals with successful immune control^12-17^.

Based on these observations, we hypothesized that T-cells induced against SARS-CoV-2 may differ in quantity and functionality depending on the severity of symptoms of COVID-19. Moreover, we hypothesized that antigen-specific T-cell characteristics may affect B-cell subpopulations and SARS-CoV-2 specific antibodies. We therefore recruited two groups of patients who were similar in the time elapsed since onset of clinical symptoms. One group included hospitalized patients with severe course of disease, whereas a second group comprised convalescent individuals who had mild disease manifestations and who completely recovered from SARS-CoV-2 related symptoms mainly in an outpatient setting.

## Results

### Study population

In this study, 50 patients with COVID-19 were included at a median of 42.5 (IQR 16.5) days after onset of symptoms. Among those, 14 were critically ill patients (64.3±8.2 years) hospitalized in the intensive care unit (“ICU patients”), whereas 36 individuals (42.2±13.6 years) had recovered from COVID-19 in an outpatient setting (“convalescent patients”) with no or mild remaining symptoms at the time of analysis (cough (n=3), rhinitis (n=2), myalgia (n=2), anosmia (n=7)). Both groups did not differ in the median time after onset of symptoms at the time of analysis (ICU patients: 40.0 (IQR 15.0) days, convalescents: 43.5 (IQR 16.5) days, p=0.37). Ten individuals without evidence for SARS-CoV-2 infection were recruited as negative controls (48.1±11.4 years). The demographic and clinical characteristics of patients and controls are shown in table 1. As expected, ICU patients were significantly older as compared to the other groups (p<0.0001). Cardiovascular disease (10/14) and metabolic diseases (7/14, especially obesity) were the most common comorbidities in ICU patients. Median time from symptom onset to hospital admission was 5 (IQR 5.5) days, and 7 (IQR 6) days to ICU admission. Eleven patients were mechanically ventilated, of which 7 were additionally treated with extracorporeal membrane oxygenation, and 7 received renal replacement therapy. Therapeutic drug regimens included hydroxychloroquine and azithromycin in 11 cases, 1 patient received tocilizumab, one patient received icatibant, and two patients underwent a three-day course of high-dose steroid treatment. Three patients died 8, 15, and 16 days after analysis, of which one still was SARS-CoV-2 PCR-positive. Twelve out of 14 ICU patients became SARS-CoV-2-PCR negative during the hospital stay. PCR results on follow-up were not available for one patient who was readmitted to the primary care hospital after end of mechanical ventilation and clinical stabilization. SARS-CoV-2 PCR was performed in 33/36 convalescents after quarantine, and all tests were negative.

**Table 1:** Demographic and clinical characteristics of the study population.

|  | Controls | SARS-CoV-2 infected |  | p-value |
| --- | --- | --- | --- | --- |
|  |  | ICU patients | convalescent<sup>a</sup> |  |
| Number | n=10 | n=14 | n=36 | n.a. |
| Years of age (mean±SD) | 48.1±11.4 | 64.3±8.2 | 42.2±13.6<sup>b</sup> | <0.0001<sup>c</sup> |
| Gender (% females) | 7/10 (70%) | 1/14 (7.1%) | 17/34 (50%)<sup>b</sup> | 0.004<sup>d</sup> |
| Time since onset of symptoms (days) | n.a. | 40 (15) | 43.5 (16.5) | 0.37<sup>e</sup> |
| Days from symptoms to hospitalization | n.a. | 5.3±3.8 | n.a. | n.a. |
| Days from symptoms to ICU (mean±SD) | n.a. | 7.8±3.3 | n.a. | n.a. |
| Leukocytes (cells/μl), median (IQR) | 6,940 (3,085) | 9,500 (9,350) | 6,615 (2,690) | 0.0005<sup>f</sup> |
| Granulocytes (cells/μl), median (IQR) | 4,043 (1,840) | 7,934 (8,511) | 4,575 (2,453) | 0.0004<sup>f</sup> |
| Monocytes (cells/μl), median (IQR) | 589 (309) | 365 (620) | 592 (226) | 0.323<sup>f</sup> |
| Lymphocytes (cells/μl), median (IQR) | 2,381 (1,349) | 910 (458) | 2,307 (728) | <0.0001<sup>f</sup> |

<sup>a</sup>Differential blood counts were available for 21/36 convalescent individuals, <sup>b</sup>Two convalescent individuals were included without knowledge of age and gender, <sup>c</sup>One-way ANOVA, <sup>d</sup> $\chi^2$ test, <sup>e</sup>Mann Whitney test, <sup>f</sup>Kruskal-Wallis test; ICU, intensive care unit; IQR, interquartile range; SD, standard deviation.

### Altered counts of leukocytes and lymphocyte subpopulations in patients with severe COVID-19

Leukocyte numbers and differential white blood counts showed substantial differences between ICU patients and convalescent individuals, with increased levels of neutrophils and severe lymphopenia as the most prominent findings (table 1). In contrast, convalescent individuals had similar levels as controls (table 1). A more detailed analysis of lymphocytes and their subpopulations was performed from whole blood using flow cytometry. Absolute cell counts were calculated based on differential blood counts. As shown in figure 1, lymphopenia affected all major lymphocyte subpopulations such as NK-cells, B-cells, and T-cells including CD4 and CD8 T-cells, and regulatory T-cells (T_reg_, data not shown).

**Figure 1:**
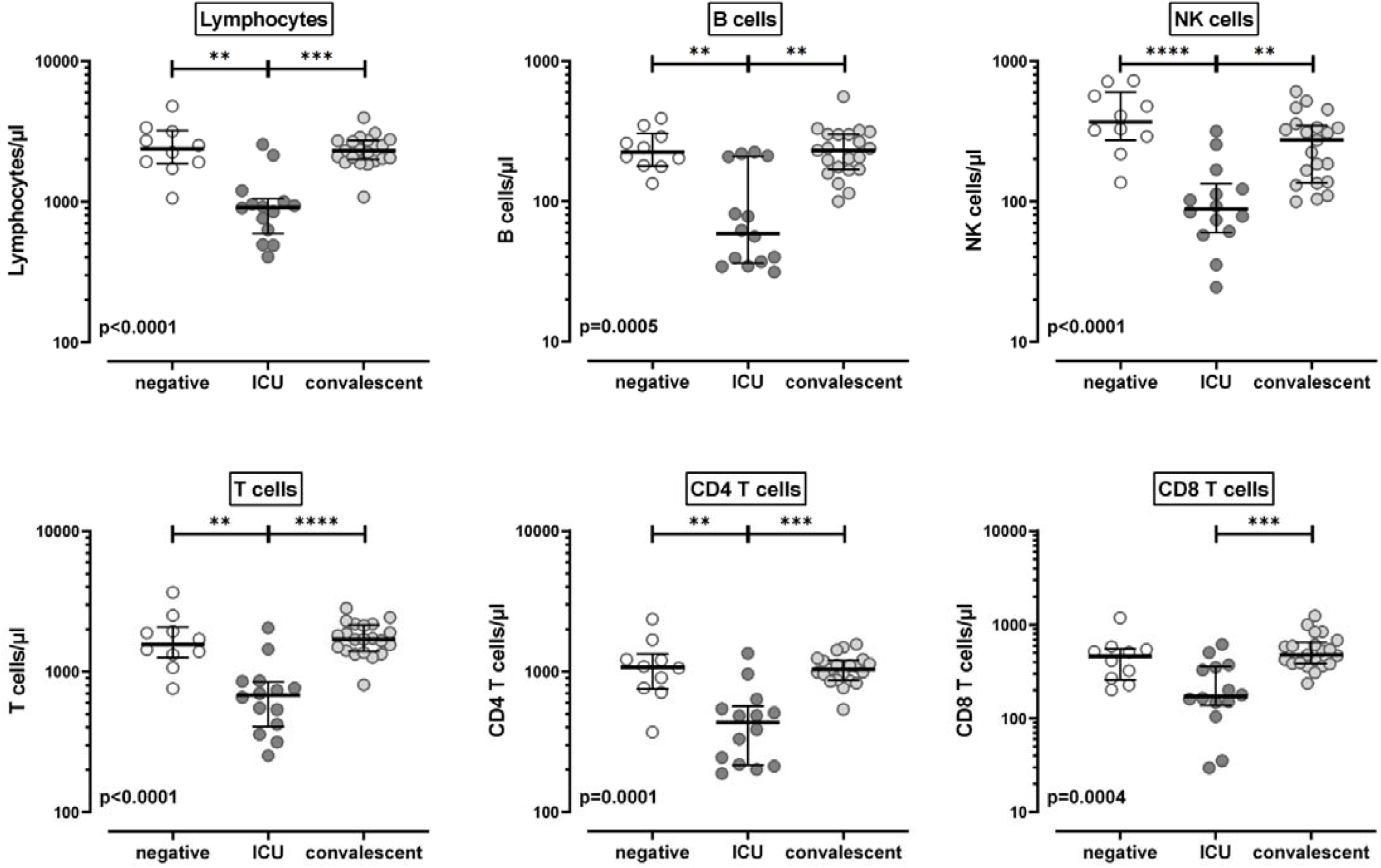
Reduced counts of lymphocytes and lymphocyte subpopulations in patients with severe COVID-19. Absolute cell numbers per µl whole blood of lymphocytes and lymphocyte subpopulations were calculated in SARS-CoV-2 negative individuals (n=10), patients with severe COVID-19 (n=14) and convalescent patients (n=21) based on flow-cytometry and differential blood counts. Flow-cytometry data were obtained from all convalescents, but 15/36 had to be excluded because no differential blood count was available. Natural killer (NK) cells were defined as CD3^−^ CD16^+^ /CD56^+^, B-cells as CD19^+^, T-cells as CD3^+^, and CD4 and CD8 T-cells as CD4^+^ CD8^−^ and CD8^+^ CD4^−^T-cells within lymphocytes, respectively. Bars represent medians with interquartile ranges. Differences between the groups were calculated using Kruskal-Wallis test and Dunn’s post test. **P<0.01, ***P<0.001, ****P<0.0001.

### Significantly higher percentages of SARS-CoV-2 specific T-cells in patients with severe COVID-19

To identify specific immunity towards SARS-CoV-2, whole blood samples were stimulated with overlapping peptide pools covering the major SARS-CoV-2 structural proteins spike (spike-N and C-terminal peptide sets, respectively), the nucleocapsid (NCAP), the membrane protein VME1, and the envelope small membrane protein VEMP. Stimulation was carried out for 6 hours and antigen-specific T-cells were identified by intracellular staining of IFNγ among activated CD69-positive CD4 and CD8 T-cells. Stimulation with *Staphylococcus aureus* Enterotoxin B (SEB) allowed assessment of polyclonal T-cell responses. DMSO was used to control for background reactivity which was subtracted from specific stimulations. A typical set of dot plots from a hospitalized patient illustrating induction of SARS-CoV-2 specific T-cell reactivity among CD4 and CD8 T-cells is shown in figure 2A. When analyzing all individuals, CD4 T-cell frequencies were highest after stimulation with spike-N and VME1, followed by spike-C and NCAP, whereas reactivity towards the smallest protein VEMP was largely absent. Antigen-specific CD8 T-cell levels were generally lower with most pronounced reactivity after stimulation with spike-N and NCAP. In contrast, spike-C, VME1, or VEMP elicited only modest or no reactivity with no difference between infected and non-infected groups (figure 2B). When comparing antigen reactivity in the three groups, significantly higher levels of SARS-CoV-2 specific CD4 T-cells were found in both infected patient groups as compared to negative controls, who were largely non-responsive (figure 2B). A difference between infected and non-infected individuals was also observed for CD8 T-cells reacting towards spike-N or NCAP, whereas CD8 T-cell reactivity towards the remaining peptide pools was equally low in both infected and in the non-infected group (figure 2B). Interestingly, among convalescents, individuals with lower respiratory symptoms such as cough or dyspnea (n=19) had significantly higher median levels of SARS-CoV-2 specific CD4 T-cells (0.16% (IQR 0.17%)) than individuals without these symptoms (n=17; 0.08% (IQR 0.127%); p=0.015; data not shown).

**Figure 2:**
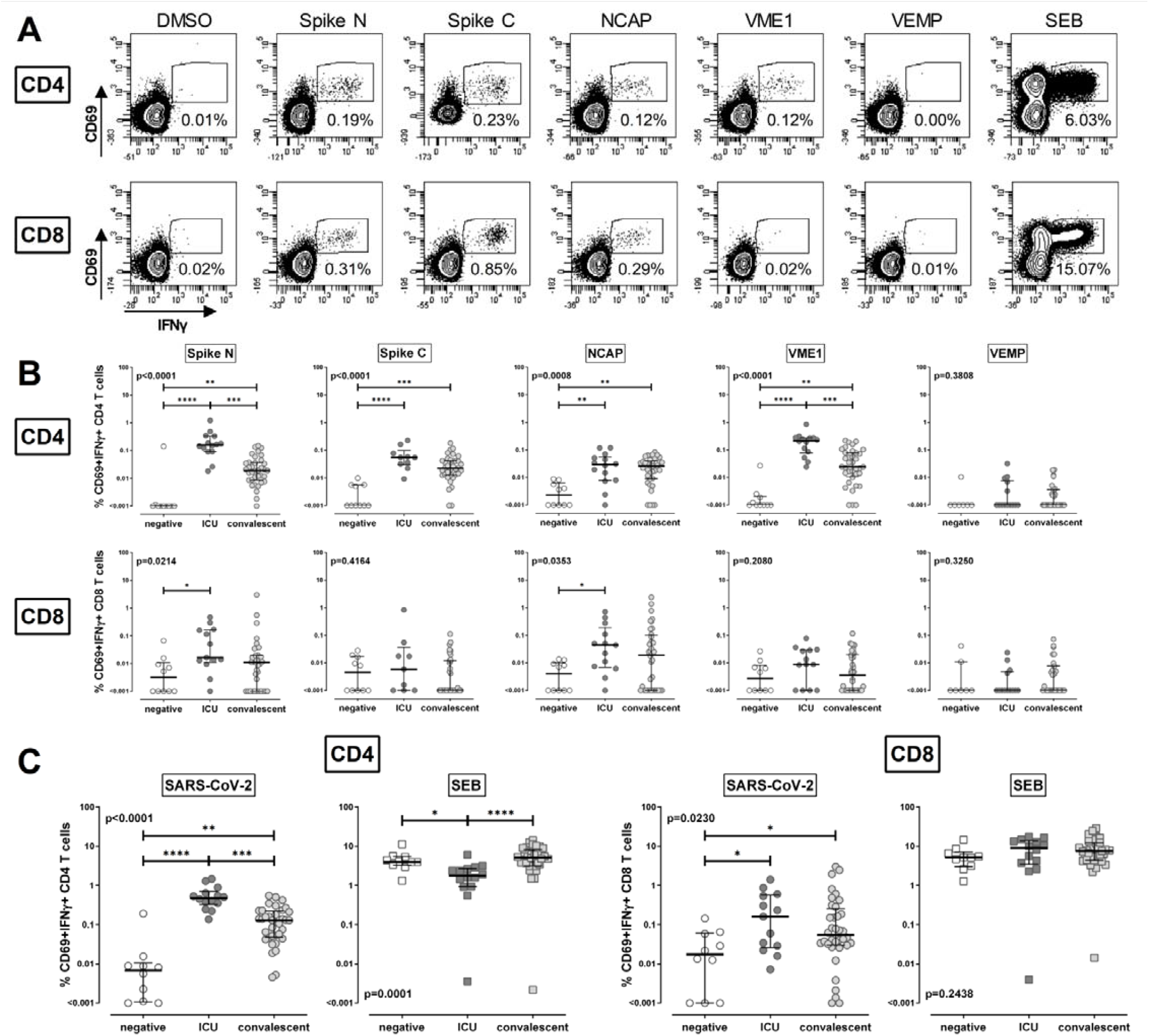
Increased percentages of SARS-CoV-2 specific T-cells in patients with severe COVID-19. Whole blood samples were stimulated with overlapping peptide pools spanning the SARS-CoV-2 spike protein (spike N, N-terminal; spike C, C-terminal), the nucleocapsid protein (NCAP), the membrane protein (VME1), and the envelope small membrane protein (VEMP). Stimulations with DMSO and *Staphylococcus aureus* enterotoxin B (SEB) served as negative controls and polyclonal stimulus, respectively. **(A)** Contour plots illustrating specific immunity from a 56-year old hospitalised patient are shown. Numbers indicate percentage of reactive (CD69 ^+^ IFNγ^+^) cells within total CD4 or CD8 T-cells. **(B)** Percentages of CD4 and CD8 T-cells specific for the different SARS-CoV-2 antigens were compared between SARS-CoV-2 negative individuals (negative, n=10), patients with severe COVID-19 (ICU, n=14) and convalescent patients (n=36). **(C)** Total percentages of SARS-CoV-2 specific T-cells, determined by the sum of frequencies towards the individual peptide pools for each individual, and SEB-reactive T-cell frequencies are compared between the three groups. Bars represent medians with interquartile ranges. Differences between the groups were calculated using Kruskal-Wallis test and Dunn’s post test. *P<0.05, **P<0.01, ***P<0.001, ****P<0.0001. IFN, interferon.

To obtain an estimate of the total levels of SARS-CoV-2 specific T-cells in each group, SARS-CoV-2 specific T-cell frequencies towards the individual peptide pools were added up for each individual (figure 2C). This showed that patients with severe course had the highest levels of SARS-CoV-2 specific CD4 T-cells (0.48% (IQR 0.37%)), which not only differed from negative controls (0.01% (IQR 0.01%)), but also from convalescent individuals (0.13% (IQR 0.18%), p<0.0001). This contrasts with polyclonal SEB-reactive CD4 T-cell frequencies, which were significantly lower in ICU patients (1.77% (IQR 1.76)) as compared to controls (3.97% (IQR 2.15)) or convalescents (5.06% (IQR 5.07), p=0.0001, figure 2C). Likewise, total levels of SARS-CoV-2 specific CD8 T-cells were significantly higher in patients than in non-infected controls (p=0.023), whereas the difference between ICU patients and convalescents did not reach statistical significance. Unlike in CD4 T-cells, SEB-reactive CD8 T-cell levels were similar among the three groups (p=0.244). SARS-CoV-2 specific CD8 T-cell levels inversely correlated with time since onset of clinical symptoms (r=-0.37, p=0.01), whereas this was not significant for specific CD4 T-cells (r=-0.24, p=0.1; data not shown). Taken together, despite strong lymphopenia affecting both CD4 and CD8 T-cells, and lower levels of polyclonal SEB-reactive CD4 T-cells, patients with severe COVID-19 were capable of mounting high levels of SARS-CoV-2 specific T-cells.

### Impaired functionality of SARS-CoV-2 specific CD4 T-cells in patients with severe course of COVID-19

To characterize the functionality of SARS-CoV-2 specific T-cells in more detail, their cytokine expression profile regarding IFNγ, IL-2 and TNFα was analyzed using flow cytometry. This resulted in assessment of a total of 7 subpopulations of cells either producing all three cytokines, two cytokines or one cytokine only (figure 3A). To ensure robust statistical analysis, this analysis was restricted to CD4 T-cells and to all samples where the total number of measurable CD69-positive IFNγ producing cells reached at least 35 (all ICU patients and 20 convalescents). As shown in figure 3A, the percentage of multifunctional SARS-CoV-2 specific CD4 T-cells producing all three cytokines was significantly lower in patients with severe course as compared to convalescent individuals. This was associated with a concomitant higher expression of cells producing IL-2 and TNFα. The overall cytokine profile of SEB-reactive CD4 T-cells was different from that of SARS-CoV-2 specific cells. Nevertheless, the SEB-reactive and SARS-CoV-2 specific cytokine profiles exhibited similar differences between patients with severe disease and convalescent individuals (figure 3A). In addition, the cytokine expression profile of SEB-reactive CD4 T-cells in convalescent individuals did not differ from SARS-CoV-2 non-infected controls (data not shown). This indicates that patients with severe disease have a restricted cytokine expression profile. Unlike in convalescents, this also extends to polyclonal T-cells in general.

**Figure 3:**
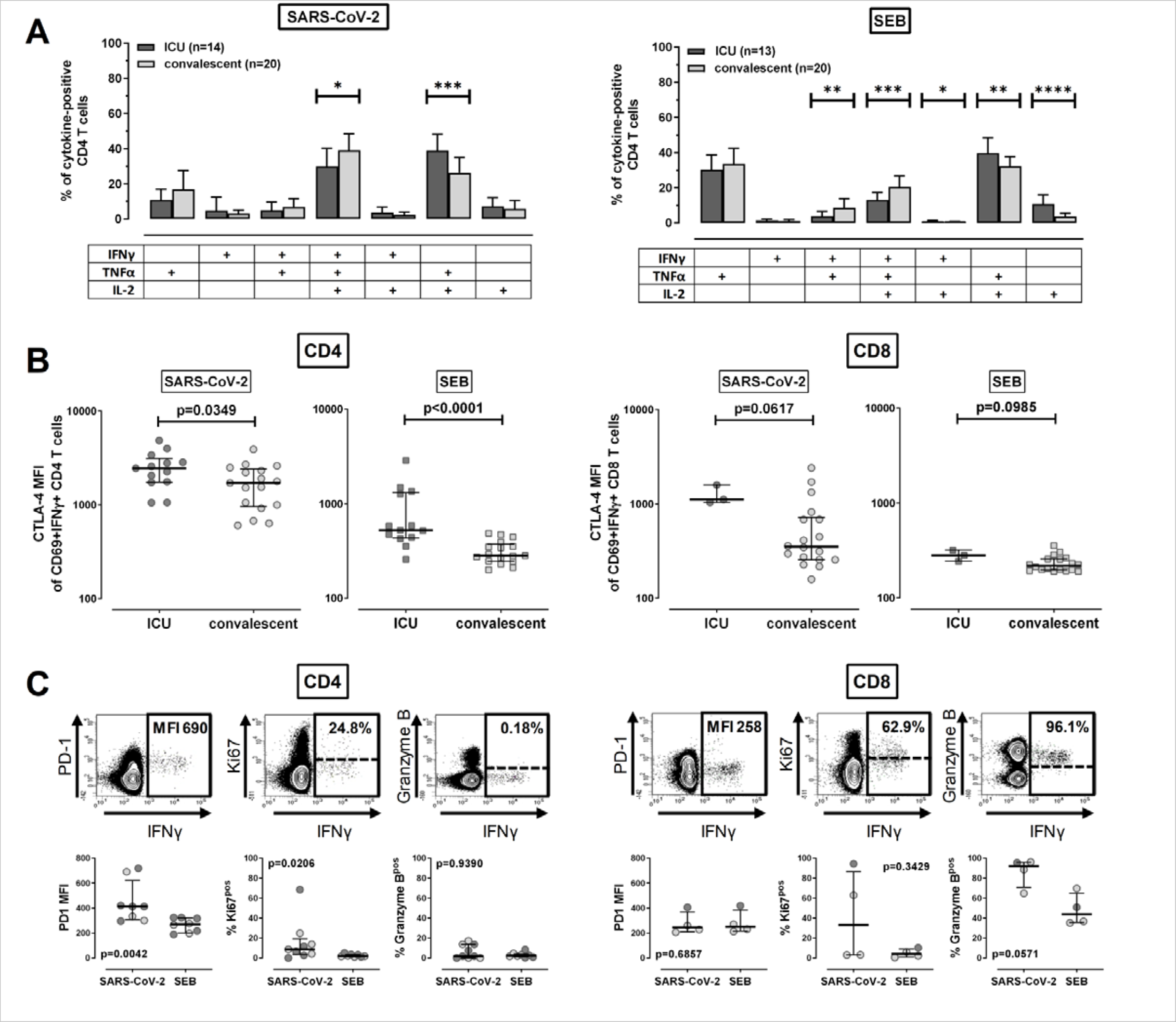
Altered cytokine profiles and phenotype of SARS-CoV-2 specific T-cells in patients with severe course of COVID-19. Expression patterns of SARS-CoV-2 specific T-cells were determined from combined T-cells reacting towards the individual peptide pools for each individual. **(A)** SARS-CoV-2 specific and *Staphylococcus* Enterotoxin B (SEB) reactive CD4 T-cells were divided into 7 subpopulations according to their expression of the cytokines IFNγ, IL-2 and TNFα. Distribution of these subgroups were compared between ICU patients and convalescents. To ensure robust statistical analysis, cytokine profiling was restricted to CD4 T-cells and to all samples with at least 35 measurable CD69^+^ IFNγ^+^ cells (all ICU patients and 20 convalescents). **(B)** CTLA-4-expression of SARS-CoV-2 specific and SEB-reactive CD4 and CD8 T-cells were compared between ICU patients and convalescents. Analysis was restricted to individuals with sufficient SARS-CoV-2 specific immunity, i.e. where the total number of measurable CD69^+^ IFNγ^+^ cells reached at least 20 cells (n=13 and 3 ICU patients and 17 and 18 convalescents for CD4 and CD8 T-cells, respectively). **(C)** In a subgroup of 10 patient samples (5 ICU patients and 5 convalescents), where a larger sample volume for in vitro stimulations was available, expression of PD-1, Ki67 and Granzyme B of SARS-CoV-2-specific and SEB-reactive CD4 and CD8 T-cells was analyzed. Overlayed contour plots (built using BD FACSdiva software 8) of samples from a 31-year old convalescent stimulated with SARS-CoV-2 antigens are shown in the upper panel. PD-1 median fluorescence intensity (MFI) was analyzed from all stimulatory reactions with at least 20 CD69^+^ IFNγ^+^ cells (n=8 and 4 for CD4 and CD8 T-cells respectively). Analysis of intranuclear presence of Ki67 (%Ki67^+^) and expression of granzyme B (%granzyme B^+^) was restricted to samples with at least 20 SARS-CoV-2 specific CD4 (n=8 for SARS-CoV-2 and n=7 for SEB) or CD8 T-cells (n=4), respectively. ICU patients are depicted by dark symbols, convalescent patients by light symbols. Bar charts in **(A)** represent mean and standard deviation and differences between the two groups were assessed using unpaired t test. Bars in **(B)** and **(C)** represent medians with interquartile ranges. Differences between the groups were calculated using Mann-Whitney test. *P<0.05, **P<0.01, ***P<0.001, ****P<0.0001. CTLA-4, cytotoxic T lymphocyte antigen 4; IFN, interferon; IL, interleukin; PD-1, programmed cell death 1; TNF tumor necrosis factor.

We also analyzed expression of CTLA-4 as phenotypical correlate of altered functionality commonly observed during active infections. This showed that SARS-CoV-2 specific CD4 T-cells from ICU patients had significantly higher expression levels of CTLA-4 than convalescents (p=0.035), which also held true for SEB-reactive CD4 T-cells (p<0.0001, figure 3B). Although the total number of patients with measurable SARS-CoV-2 reactive CD8 T-cells was lower, a similar trend was found for SARS-CoV-2 reactive or SEB-reactive CD8 T-cells (figure 3B).

Finally, a subset of 10 SARS-CoV-2 infected patients (5 hospitalized, 5 convalescents) was studied to further characterize SARS-CoV-2 specific CD4 and CD8 T-cells for expression of PD-1, Ki67, and granzyme B, with dot plots shown in figure 3C. PD-1 expression levels and the percentage of Ki67 positive cells were higher on SARS-CoV-2 specific CD4 T-cells than on polyclonal SEB-reactive CD4 T-cells. Likewise, some individuals had Ki67 expressing CD8 T-cells, and a large fraction of SARS-CoV-2 specific CD8 T-cells expressed granzyme B which was lower among SEB-reactive T-cells (figure 3C).

### Altered phenotype of global CD4 and CD8 T-cells in patients with severe COVID-19

As cytokine expression patterns and CTLA-4 expression in patients with severe course was altered in both SARS-CoV-2 specific and SEB-reactive T-cells, these alterations may also extend to T-cells in general. To analyze phenotypical characteristics of bulk T-cells in more detail, expression of CTLA-4 and PD-1, as well as Ki67 positive cells were analyzed directly from whole blood without prior stimulation. As shown in figure 4, both CD4 and CD8 T-cells from ICU patients showed markedly increased expression of CTLA-4 and PD-1 as compared to controls, whereas respective expression in convalescent individuals was lower and similar as in controls. Interestingly, the percentage of recently proliferated Ki67 positive CD4 and CD8 T-cells was significantly higher in patients with severe course as compared to controls and convalescent individuals.

**Figure 4:**
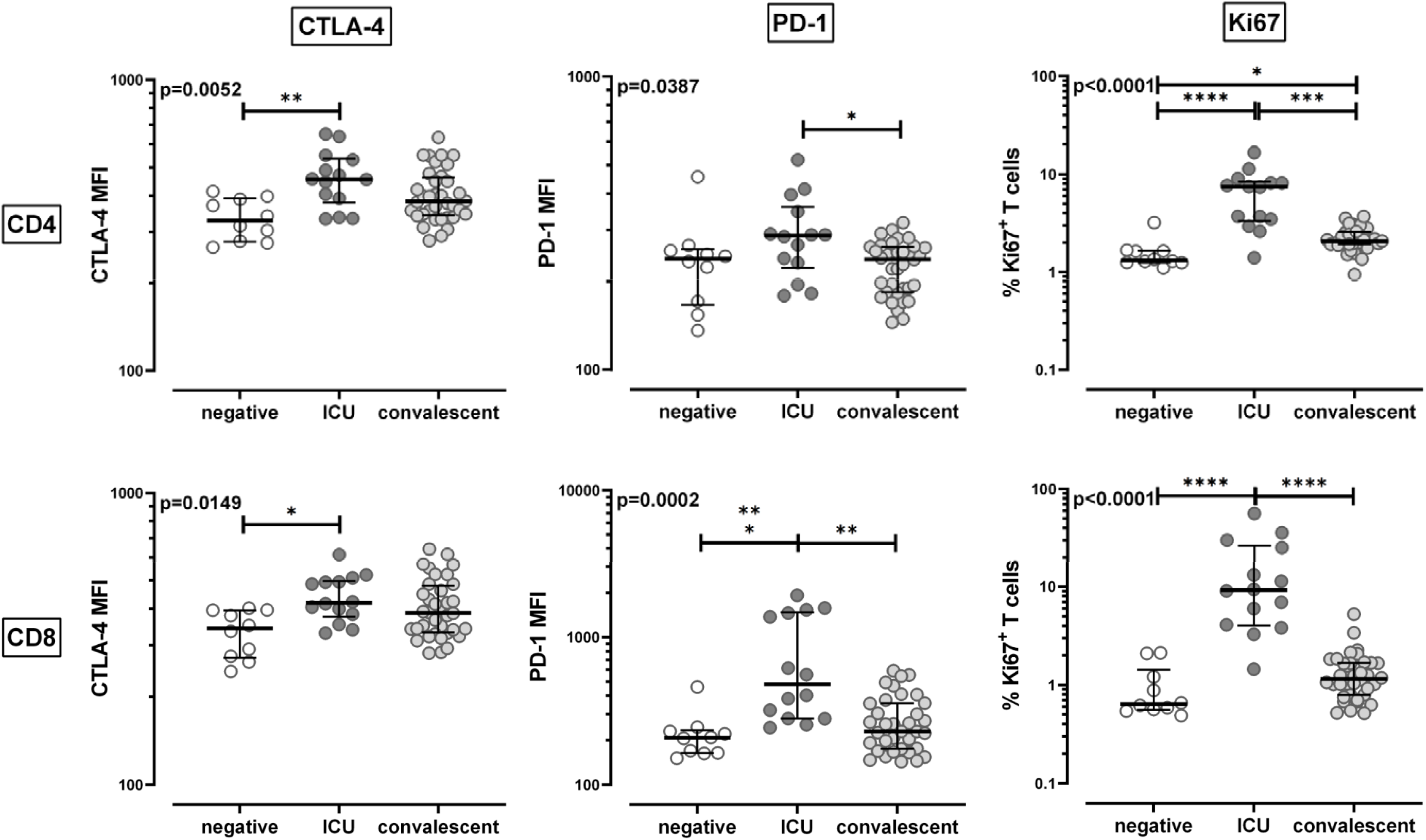
Altered phenotype of global CD4 and CD8 T-cells in patients with severe COVID-19. Expression of cytotoxic T lymphocyte antigen 4 (CTLA-4), programmed cell death 1 (PD-1) and intranuclear Ki67-expression of unstimulated total CD4 and CD8 T-cells were compared between SARS-CoV-2 negative individuals (negative, n=10), patients with severe COVID-19 (ICU, n=14) and convalescent patients (n=36). Bars represent medians with interquartile ranges. Differences between the groups were calculated using Kruskal-Wallis test and Dunn’s post test. *P<0.05, **P<0.01, ***P<0.001, ****P<0.0001.

### Strong correlation of SARS-CoV-2 specific CD4 T-cell levels with specific IgG and IgA antibodies and plasmablasts

To comparatively analyze cellular and humoral immunity towards SARS-CoV-2, specific IgA and IgG antibodies were determined using ELISA. As shown in figure 5A, all individuals with severe course were positive for SARS-CoV-2 specific IgG and IgA. Interestingly, their levels were significantly higher than those of convalescent individuals where only 83% (30/36) had positive IgG and 69% (25/36) had IgA above detection limit. Intermediate IgA and IgG titers were found in two individuals each. SARS-CoV-2 negative controls did not show any specific IgG or IgA. In line with the role of CD4 T-cells in providing help for induction of humoral immunity, the percentage of SARS-CoV-2 specific CD4 T-cells showed a significant correlation with both specific IgG (r=0.77, p<0.0001) and IgA antibodies (r=0.67, p<0.0001), whereas no correlation was observed with specific CD8 T-cells (p=0.78 for IgG, p=0.52 for IgA, figure 5B). To elucidate whether the observed differences in specific antibody levels were related to differences in B-cells among the groups, we analyzed CD19 positive B-cell subpopulations by their expression of IgD and CD27, with dot plots of a 64 years old hospitalized patient shown in figure 5C. As with B-cell lymphopenia in general (figure 1), the numbers of naïve (IgD+/CD27-), non-switched memory (IgD+/CD27+) and switched memory B-cells (IgD- /CD27+) were significantly lower in patients with severe course (figure 5C). Interestingly, however, their number of plasmablasts, which were identified as CD38 positive switched memory B-cells, was significantly higher than in controls or convalescents. In line with the central role of plasmablasts in initiating antibody production, their numbers showed a strong correlation with both IgG (r=0.53, p=0.0014) and IgA antibody levels (r=0.54, p=0.0013, figure 5D).

**Figure 5:**
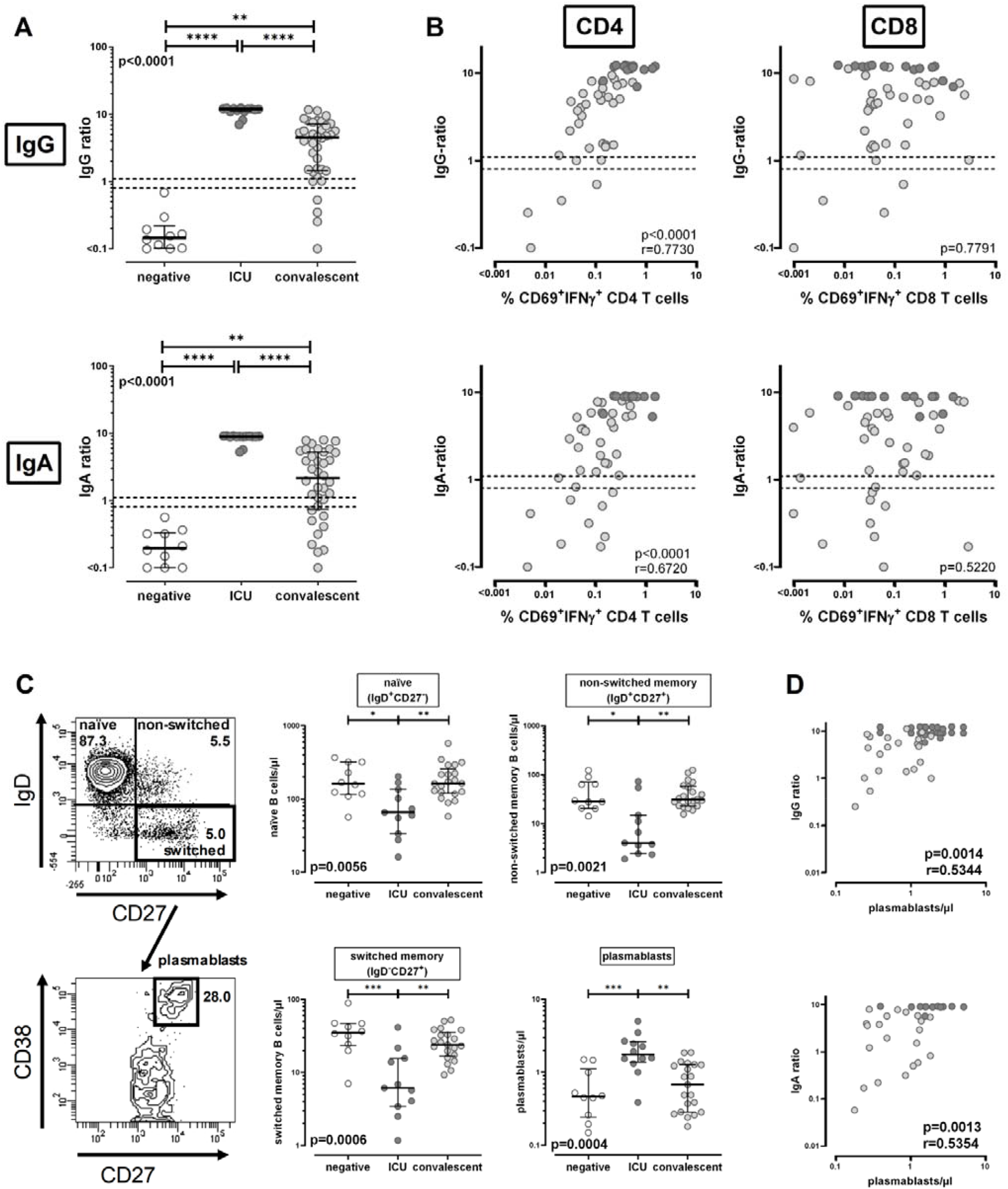
Strong correlation of SARS-CoV-2 specific CD4 T-cell levels with specific IgG and IgA antibodies and plasmablasts. **(A)** Levels of SARS-CoV-2 specific IgG and IgA were compared between SARS-CoV-2 negative individuals (negative, n=10), patients with severe COVID-19 (ICU, n=14) and convalescent patients (n=36). **(B)** Correlation between levels of SARS-CoV-2 specific IgG or IgA with frequencies of SARS-CoV-2 specific CD4 or CD8 T-cells in patients with SARS-CoV-2 infected individuals. **(C)** Representative contour plots of a 64-years old hospitalized patient showing the differentiation status of B-cells characterized by surface expression of IgD and CD27, with plasmablasts identified among switched-memory B-cells by additional staining of CD38. Numbers of B-cell subpopulations and plasmablasts were compared between groups and **(D)** plasmablasts were correlated with levels of SARS-CoV-2 specific IgG and IgA. Antibody levels were determined semiquantitatively by dividing the optical density of an individual sample by that of a positive control serum. Bars in (A) and (C) represent medians with interquartile ranges. Differences between the groups were calculated using Kruskal-Wallis test and Dunn’s post test. *P<0.05, **P<0.01, ***P<0.001, ****P<0.0001. Correlations in (B) and (D) were analyzed according to Spearman. Dotted lines indicate detection limits for IgG and IgA, indicating negative, intermediate and positive levels, respectively as per manufacturer’s instructions. IFN, interferon.

## Discussion

Manifestation of SARS-CoV-2 infections may range from asymptomatic infections or mild symptoms to severe courses of disease with a high risk of fatal outcome^2^. In this study, we show that SARS-CoV-2 specific immunological characteristics in patients with severe course are clearly distinct from infected individuals who recovered from mild disease that could be managed in an outpatient setting. Both groups were analyzed at the same time after onset of COVID-19 symptoms. As main findings, we show that patients with severe disease had high levels of SARS-CoV-2 specific CD4 and CD8 T-cells as well as high titers of specific IgG and IgA antibodies as compared to convalescent individuals, where levels were significantly lower. However, SARS-CoV-2 specific T-cells in severe cases had a restricted cytokine expression profile with less multifunctional cells, and strongly expressed CTLA-4 as hallmark of T-cells in the contraction phase of an immune response after active encounter with the virus. In contrast, convalescent individuals who had recovered from mild or moderate disease had lower levels of SARS-CoV-2 specific humoral and cellular immunity, and antigen-specific T-cells showed less signs of functional alterations. Finally, quantitative and phenotypical differences between the infected patient groups were also found for major lymphocyte subpopulations such as B-cells, NK cells, Treg, and CD4 and CD8 T-cells. Apart from severe lymphopenia, CD4 and CD8 T-cells from patients with severe COVID-19 exhibited increased expression of CTLA-4 and PD-1, and a high expression of Ki67 as marker for recent proliferation. In addition, the percentage of cells responding after polyclonal stimulation was lower and restricted in functionality. In contrast, lymphocyte characteristics from convalescent individuals were similar as in non-infected controls. Taken together, this indicates that the severity of clinical disease in patients with COVID-19 is not only associated with prominent changes in the innate immune system but is also characterized by a marked alteration of adaptive humoral and cellular immunity that includes both SARS-CoV-2 specific and global T-cell function.

Up to now, only few studies have described SARS-CoV-2 specific T-cells in patients with COVID-19^7, 8, 18, 19^. Together with results from our study, key characteristics of SARS-CoV-2 specific T-cells emerge. SARS-CoV-2 reactive T-cells exhibit immediate effector function with proliferative potential, and expression of IFNγ, IL-2 and TNFα, which suggests a Th1 phenotype. This is supported by results from supernatants of stimulated peripheral blood mononuclear cells showing detectable IFNγ, IL-2 and TNFα, and low levels of IL-5, IL-13, IL-9, IL-10, and IL-22^7, 18^. Reactive T-cells exist among both CD4 and CD8 T-cells, and CD8 T-cells were able to produce effector molecules such as granzyme B. In line with two other reports^8, 18^, SARS-CoV-2 specific CD4 T-cell levels were higher than those of CD8 T-cells. Unlike CD4 T-cells, specific CD8 T-cell levels inversely correlated with time after onset of symptoms, which may reflect higher stability of CD4 T-cells. Alternatively, antigen-specific CD8 T-cells might have been recruited to the lungs as the site of massive SARS-CoV-2 replication, as signatures for clonally expanded CD8 T-cells were found in bronchoalveolar lavage samples of patients with SARS-CoV-2 associated lung disease^20^. All studies show that SARS-CoV-2 specific T-cell levels differed in infected and non-infected individuals^7, 8, 18, 19^. Our results suggest that SARS-CoV-2 specific CD4 T-cells allowed for a better distinction not only of non-infected and infected individuals but also of patients with different severity of disease. This distinction may further be improved using immunodominant peptides for stimulation. So far, a relative immunodominance of the spike protein was described^8, 18, 19^. This was also observed in our study, but the four viral structural proteins differed in their ability to induce specific immunity among CD4 and CD8 T-cells. The VEMP protein hardly elicited any reactivity, which may be related to its small size and its relatively low abundance in the virus particle^21^. Reactive CD4 T-cells were found towards all other proteins with a dominance of the spike and the VME1 protein. Interestingly, apart from the N-terminal portion of the spike protein, CD8 T-cells showed pronounced reactivity towards the NCAP protein. NCAP may be more readily processed to be presented in MHC class I molecules due to its predominant localization in the cytoplasm, whereas all other membrane proteins are directly assembled in the ER membrane^22^.

It was striking that patients with severe course of disease had significantly higher levels of both antibodies and T-cells as compared to convalescents. As all analyses were performed in a short time frame after onset of symptoms, it is considered unlikely that antibody and T-cell levels in convalescent individuals had been similarly high during active viral replication and had decreased after successful control of infection. Instead, the levels of specific humoral and cellular immunity needed to control viral replication may be directly related to the viral load during primary infection. Thus, patients with severe course may have required induction of higher levels of specific immunity due to higher viral load or prolonged periods of active viral replication. Infection efficiency is high in nasal epithelial cells of the upper airways, and decreases in epithelial cells of the lower respiratory tract along an angiotensin converting enzyme 2 (ACE2) receptor gradient^23^. Therefore, viral replication may remain restricted to the upper airway in the majority of infected individuals with mild symptoms. Further seeding of virus to the lung may be favored by high viral load in the upper airways with subsequent microaspiration events that are more frequent in patients at risk for severe courses of COVID-19 such as the elderly, diabetic or obese individuals. Thus, lower viral load with local restriction to the upper airways may require less pronounced specific immunity as compared to higher viral load and/or further dissemination of the virus to the lower respiratory tract. This may be supported by our observation that SARS-CoV-2 specific CD4 T-cell levels showed significant differences among convalescent individuals with or without symptoms of the lower respiratory tract (cough and dyspnea). The Induction of specific immunity may further be modulated by pre-existing cross-reactive immunity towards common cold coronaviruses. This is illustrated by influenza-vaccine studies, were pre-existing immunity towards influenza is associated with a less pronounced induction of vaccine-specific immunity as compared to influenza-naïve subjects^24, 25^. Evidence for cross-reactive immunity also exists among coronaviruses. In our study, very low levels of SARS-CoV-2 reactive T-cells were in part detectable among control subjects without SARS-CoV-2 infection. However, as shown in recent studies using longer stimulation times, evidence for cross-reactive T-cells was found in 20 to up to 50% of non-infected controls^8, 18^.

Based on a variety of clinically relevant pathogens, the quantity and the phenotype of antigen-specific T-cells has been shown to differ in relation to the pathogen activity in the context of primary infections or reactivations. As exemplified for immunity towards cytomegalovirus (CMV), varicella zoster virus (VZV), human immunodeficiency virus (HIV), or mycobacteria, T-cells induced by primary infection or reactivation during active encounter with the pathogen show a low percentage of multifunctional cells and increased expression of inhibitory surface receptors such as CTLA-4 or PD-1, whereas the expression of these molecules decreases with successful control of the pathogen^12-15, 26, 27^. In this respect, the lower CTLA-4 expression levels on SARS-CoV-2 specific T-cells of convalescent individuals are compatible with successful viral control, whereas the increased expression of CTLA-4 on SARS-CoV-2 specific T-cells in patients with severe disease is in line with a prolonged and more intense encounter with the virus. Consistent with primary induction, specific T-cells had a restricted cytokine pattern with a low percentage of multifunctional cells and a relative dominance of single or dual cytokine-producing cells expressing IL-2, which is different from reactivations, where the loss in multifunctional cells is associated with a shift towards cells exclusively expressing IFNγ^15, 28^. Although this functional profile of SARS-CoV-2 specific T-cells in patients with severe disease has several characteristics of an exhausted phenotype found in patients with symptomatic disease in the context of chronic infections and/or reactivations, exhaustion is frequently associated with a quantitative decrease in specific T-cells^12, 14^. In contrast, our patients were able to mount a strong adaptive T-cell response with proliferative potential, and the majority of patients achieved control of viral replication. Therefore, the high expression levels of CTLA-4 and the restricted functionality may likely reflect a physiological contraction mechanism to downregulate specific immunity after its strong induction and to compensate for excessive immunopathology in the lung. This process appears to have notable effects on lymphocyte subpopulations and their functional characteristics in general, which show the same pattern of inhibitory surface receptors and functional restriction, and thereby may account for an increased susceptibility for other opportunistic infections in patients with severe COVID-19^29, 30^.

As with SARS-CoV-2 specific T-cells, specific antibody responses were also highest in patients with severe disease. Interestingly, despite severe B-cell lymphopenia which affected all major subpopulations, the increased antibody levels showed a direct correlation with the number of circulating plasmablasts, which were significantly higher than in convalescents and non-infected controls. As high antibody responses were shown to correlate with neutralization capacity, this may directly contribute to viral clearance^10, 11, 31^. However, given the association with disease severity, further studies should address whether antibodies may also contribute antibody-dependent enhancement of viral entry into Fc-receptor expressing cells such as macrophages thereby leading to increased inflammation and lung injury^32^.

Our study is limited by a low sample size. Nevertheless, differences in general as well as antigen-specific immunity between the two patient groups are very pronounced and correlate well with the severity of the disease. Moreover, we did not perform any longitudinal analyses of specific T-cells and antibodies to evaluate whether the levels of specific immunity during primary infection will determine stability and protection in the long-term. Data on SARS-CoV-1 specific immunity indicated that both antibodies and T-cells were detectable for several years, with highest stability in patients with more severe disease^33, 34^. Similar studies with larger sample size are needed to evaluate whether the more pronounced immunity in patients with severe COVID-19 may result in higher stability and better protection from reinfection with SARS-CoV-2 in the long-term.

Knowledge gained from this study may have implications for vaccine design and therapeutic management. Our study revealed an immunodominance of specific T-cells towards the spike protein as the main vaccine target. In addition, other viral proteins may represent promising antigens to achieve a broad vaccine-induced T-cell response comparable with natural infection. Up to now, the role of immunosuppressive drugs for treatment of COVID-19 is controversially discussed^35^. Our results show that patients with severe disease mount a particularly strong cellular and humoral immune response. While this immune response seems to be efficient in controlling viremia, contraction is required to prevent immunopathology associated with a hyperactive immune system. It is therefore tempting to speculate whether immunosuppressive drugs are harmful when given in the induction phase but may have particular benefit in the contraction phase of the immune response. Data emerging from the Recovery Trial indeed provide first evidence for a particular survival benefit of steroid treatment in ventilated patients with severe disease (https://www.recoverytrial.net/results).

## Methods

### Study design and patient population

Patients who were hospitalized with PCR-confirmed COVID-19 (ICU patients) and patients with milder course of disease in an outpatient setting were recruited who had been matched with ICU patients according to the time since onset of clinical symptoms. In addition, individuals without evidence for infection with SARS-CoV-2 were tested as negative controls. ICU patients were recruited within the CORSAAR study, a cohort study on patients with COVID-19. The study was approved by the ethics committee of the Ärztekammer des Saarlandes (references 76/20; l62/20) and all individuals or their legal representatives gave written informed consent. Information on clinical symptoms were derived from patient charts or collected based on a questionnaire. Blood samples (4.7ml) were collected in lithium heparin containing tubes, and all analyses of antigen-specific T-cells and lymphocyte subpopulations were carried out within 24 hours. Antibody testing was performed from frozen plasma samples.

### Quantitation of lymphocyte populations

Quantitation and characterization of lymphocyte subpopulations was performed on 100 µL of heparinized whole blood as described before ^36^ using monoclonal antibodies towards CD3 (clone SK7), CD4 (clone SK3), CD8 (clone RPA-T8 and SK1), CD16 (clone 3G8), CD19 (clone HIB19), CD27 (clone L128), CD38 (clone HB7), CD56 (clone B159), cytotoxic T lymphocyte antigen 4 (CTLA-4, clone BNI3), IgD (clone IA6-2) and programmed cell death 1 (PD-1, clone MIH4, all from BD Biosciences). For samples that included anti-CD27 and anti-IgD, whole blood was washed with medium (RPMI) prior to staining to remove soluble CD27 and IgD. After 25 min of incubation, samples were treated with lysing solution (BD Biosciences). Thereafter, cells were washed with FACS-buffer (PBS, 5% filtered FCS, 0.5% bovine serum albumin, 0.07% NaN_3_), and analyzed using flow cytometry (FACS-Canto-II) and FACS-Diva-V6.1.3-software (BD Biosciences). Gating strategies for each staining procedure are provided in supplementary figure S1. Intranuclear staining of Ki67 (clone B56) was performed using the Foxp3/transcription factor staining buffer set according to the manufacturer’s instructions (eBioscience/Thermofisher). Differentiation status of CD19-positive B-cells was assessed using antibodies against IgD and CD27. Plasmablasts were identified among switched-memory B-cells by additional staining of CD38. In addition, T-cells were phenotypically analyzed for expression of PD-1 and CTLA-4. Differential blood counts were used to calculate absolute lymphocyte numbers. CD4 and CD8 T-cells were quantified among CD3 T-cells and these among lymphocytes. NK cells were identified using antibodies towards CD3, CD16 and CD56 and quantified as CD3 negative/CD16/CD56 positive lymphocytes. Detailed information on antibodies for flow cytometric stainings are given in supplementary table S1.

### Stimulation assays

Whole blood samples were stimulated with overlapping peptide pools spanning the SARS-CoV-2 spike protein (spike vial 1, N-terminal receptor binding domain and spike vial 2, C-terminal portion including the transmembrane domain), the nucleocapsid protein NCAP, the membrane protein VME1, and the envelope small membrane protein VEMP (1µg/ml each; JPT, Berlin, Germany) to induce antigen-specific activation and cytokine induction as described previously ^14^. As a negative control, samples were treated with the diluent DMSO. Cells were stimulated with 2.5 μg/mL Staphylococcus aureus enterotoxin B (SEB; Sigma) to assess general characteristics of polyclonally stimulated T-cells. Stimulation was performed from whole blood for 6 h, with 10µg/mL brefeldin A added after 2 h of incubation. After 6 h, samples were treated with 20mM EDTA for 15 min; thereafter, cells were fixed using BD lysing solution, and stimulated cells were immunostained using anti-CD4 (clone SK3), anti-CD69 (clone L78), anti-IFNγ (clone 4S.B3), anti-IL2 (clone MQ1-17H12), anti-TNFα (clone MAb11), anti-PD-1 (clone MIH4), anti-CTLA4 (clone BNI3), anti-Ki67 (clone B56), or anti-granzyme B (clone GB11). All stainings except for PD-1 were performed after fixation. Ki67 staining was performed using the Foxp3/transcription factor staining buffer set as described above. Cells were analyzed using flow-cytometry. Gating strategies are provided in supplementary figures S2 and S3.

### Analysis of SARS-CoV-2 specific antibodies

SARS-CoV-2 specific antibodies were quantified from heparinized plasma samples using an IgG and IgA assay coated with recombinant S1-domain of SARS-CoV-2 spike protein antigen according to the manufacturer’s instructions (Euroimmun, Lübeck, Germany). Antibody levels are expressed as ratios that are defined as the extinction of the patient sample divided by the extinction of a calibrator serum. Ratios <0.8 were scored negative, ratios between ≥0.8 and <1.1 were scored intermediate, and ratios ≥1.1 were scored positive.

### Statistical analysis

Statistical analysis was carried out using GraphPad Prism 8.0 software using two-tailed tests. An unpaired non-parametric Kruskall-Wallis test with Dunn’s post test was used to analyze differences for lymphocyte subpopulations, T-cell and antibody levels as wells as PD-1, CTLA-4 and Ki67 of total T-cells among the three groups. Mann-Whitney test was performed to compare non-parametric data between two groups (time since onset of symptoms, expression of CTLA-4, PD1, Ki67 and Granzyme B of specific T-cells). Data with normal distribution were analyzed using unpaired t test (cytokine expression) or one-way analysis of variance test (age). Differences in gender were analyzed using χ^2^ test. Correlations between T-cell levels, antibody titers, plasmablasts and time from onset of symptoms were analyzed according to Spearman. A p-value of <0.05 was considered statistically significant.

### Data availability

All figures have associated raw data. The data that support the findings of this study are available from the corresponding author upon reasonable request.

## Data Availability

The data that support the findings of this study are available from the corresponding author upon reasonable request.

## Acknowledgements

The authors thank Candida Guckelmus and Rebecca Urschel for excellent technical assistance. The authors also thank all participants to this study who contributed to the gain in knowledge from this project. The study was supported by institutional funds, and in part by grants of Saarland University, the State of Saarland, and the Dr. Rolf M. Schwiete Stiftung to R.B.

## Author Contributions

D.S., T.S., U.S., S.S., B.C.G., S.L.B. and M.S. designed the study; D.S., T.S., U.S. and M.S. designed the experiments, D.S., V.K., and T.S. performed experiments; S.S., B.C.G., P.M.L., H.W., R.B., J.M. and H.E. contributed to study design, patient recruitment, and clinical data acquisition. D.S., T.S., U.S. and M.S. supervised all parts of the study, performed analyses and wrote the manuscript. All authors approved the final version of the manuscript.

## Declaration of Interests

The authors declare no competing interests.

## Notes

### Competing Interest Statement

The authors have declared no competing interest.

### Author Declarations

Aerztekammer des Saarlandes (reference numbers 76/20; l62/20)

